# The New Therapeutics in Alzheimer’s Disease Longitudinal Cohort study (NTAD): study protocol

**DOI:** 10.1101/2021.05.18.21257340

**Authors:** Juliette H Lanskey, Ece Kocagoncu, Andrew J Quin, Yun-Ju Cheng, Melek Karadag, Jemma Pitt, John Isaac, Stephen Lowe, Michael Perkinton, Vanessa Raymont, Krish D Singh, Mark Woolrich, Anna C Nobre, Richard N Henson, James B Rowe, on behalf of the NTAD study group

**Affiliations:** MRC Cognition and Brain Sciences Unit, University of Cambridge, Cambridge, UK; Department of Clinical Neurosciences and Cambridge University Hospitals NHS Foundation Trust, Cambridge Biomedical Campus, Cambridge, UK; Oxford Centre for Human Brain Activity, Wellcome Centre for Integrative Neuroimaging, Department of Psychiatry, University of Oxford, Oxford, UK; Lilly Corporate Center, Indianapolis, USA; Neuroscience External Innovation, Johnson & Johnson Innovations, London, UK; Neuroscience, BioPharmaceuticals R&D, AstraZeneca, Cambridge, UK; Department of Psychiatry, University of Oxford, Oxford, UK; Cardiff University Brain Research Imaging Centre, School of Psychology, Cardiff University, Cardiff, UK; Department of Psychiatry, University of Cambridge, Cambridge, UK

## Abstract

**Introduction:** With the pressing need to develop treatments that slow or stop the progression of Alzheimer’s disease, new tools are needed to reduce clinical trial duration and validate new targets for human therapeutics. Such tools could be derived from neurophysiological measurements of disease.

**Methods and Analysis:** The New Therapeutics in Alzheimer’s disease study (NTAD) aims to identify a biomarker set from magneto/electro-encephalographic that is sensitive to disease and progression over one year. The study will recruit 100 people with amyloid-positive mild cognitive impairment or early-stage Alzheimer’s disease and 30 healthy controls aged between 50 and 85 years. Repeat measurements of the clinical, cognitive and imaging data (magnetoencephalography, electroencephalography and magnetic resonance imaging) of participants with Alzheimer’s disease or mild cognitive impairment will be taken at baseline and at one year. To assess reliability of magneto/electro-encephalographic changes, a subset of 30 participants with mild cognitive impairment or early-stage Alzheimer’s disease will undergo repeat magneto/electro-encephalographic two weeks after baseline. Clinical and cognitive assessment will be repeated at 2 years. Linear mixed models of baseline and longitudinal change in neurophysiology are the primary analyses of interest, supported by Bayesian inference. Additional outputs will include relative effect sizes for physiological markers, atrophy and cognitive change and the respective numbers needed to treat each arm of simulated clinical trials of a future disease modifying therapy.

**Ethics and dissemination:** The study has received a favourable opinion from the East of England Cambridge Central Research Ethics Committee (REC reference 18/EE/0042). Results will be disseminated through internal reports, peer-reviewed scientific journals, conference presentations, website publication, submission to regulatory authorities and other publications. Data will be made available via the Dementias Platform UK Data Portal on completion of initial analyses by the NTAD study group.

**STRENGTHS AND LIMITATIONS OF THIS STUDY:** *Strengths:*

- NTAD is a longitudinal, multicentre study of magneto/electro-encephalographic measures of Alzheimer’s disease progression
- NTAD assesses the test-retest reliability of magneto/electro-encephalographic parameters
- All participants with early-stage Alzheimer’s disease or mild cognitive impairment will be amyloid-positive

*Limitations:*

- Attrition during follow up may limit inferences
- Recruitment is from volunteer panels and clinical services that may not reflect national diversity of people affected by Alzheimer’s disease

## INTRODUCTION

With 44 million people living worldwide with dementia, treatments to slow or stop disease progression are urgently needed. Alzheimer’s disease is the most prevalent dementia but despite rapid advances in therapeutics within preclinical models,[1,2] clinical trials remain expensive, slow and challenging[3–5] with high-profile failures.[6] Significant bottlenecks exist in early-phase trials, where efficacy rates are very low and costs rapidly escalate. To bridge the gap between animal models and the human disease, better tools are needed to quantify pathogenic and pathophysiological mechanisms in patients, *in vivo* providing evidence to pursue or discontinue trials of a candidate treatment. These tools should be safe, scalable and able to support early-phase trials over a short duration and viable budget.

Brain imaging is widely used to diagnose dementia and measure pathology in clinical trials. There are diverse imaging methods with differential sensitivity to brain structure, chemistry, pathology and function. For example, magnetic resonance imaging (MRI) is commonly used to measure structural changes related to Alzheimer’s disease,[7] such as entorhinal cortex[8] and hippocampus volumes.[9] Positron emission tomography (PET) can quantify and localise Alzheimer pathology, through the use of ligands that bind to the aggregated tau protein,[10–14] beta-amyloid plaques[15] and neuroinflammation.[16]

The loss of synapses and synaptic plasticity is an early feature of Alzheimer’s disease.[17–19] Indeed, measures of synaptic loss may be more sensitive than measures of atrophy to cognitive decline.[20,21] As synaptic changes occur early in Alzheimer’s disease,[20] such measures could be sensitive to the earliest precursors of cognitive involvement. This accords with preclinical models in transgenic mice where network physiology and cognition are impaired before tangles or cell death.[22]

In contrast to preclinical models and post-mortem analysis, there are limited options to assess synaptic function in humans, *in vivo*. While PET imaging now offers ligands that indirectly quantify synaptic density, electroencephalography (EEG) and magnetoencephalography (MEG) measure neurophysiological properties that depend on synaptic integrity and function within local and large-scale brain networks. MEG can identify synaptic and local circuit impairments[23,24] and their impact on network dynamics in Alzheimer’s Disease,[25–28] frontotemporal dementia,[24,29–32] and Lewy-body disease.[33] MEG and EEG therefore have potential to support and de-risk clinical trials of novel compounds. However, to enable MEG as a viable biomarker in clinical trials, one must assess longitudinal MEG and test-retest reliability and harmonise MEG protocols across sites.

The New Therapeutics for Alzheimer’s Disease (NTAD) study aims to identify viable MEG and EEG biomarkers for clinical trials. The NTAD consortium is a multicentre study established by the Dementias Platform UK and supported by the Medical Research Council, Alzheimer’s Research UK and industry partners.

This paper describes the NTAD study and its protocol to acquire longitudinal MEG and EEG data in people with biomarker-positive mild cognitive impairment and early Alzheimer’s disease.

### Research Aims

The primary objective is to identify a biomarker set from neurophysiological MEG and EEG that is sensitive to the presence and progression of Alzheimer’s disease. We aim to harmonise MEG protocols across sites to allow biomarker identification from pooled data. Such a neurophysiological biomarker, or biomarker set, should be related to cognitive function, have high test-retest reliability and be able to track disease progression over the duration of clinical trials. The sensitivity to disease progression should ideally also outperform current widely used biomarkers such as MRI and cognitive tests.

Our secondary objective is to identify a biomarker set that can predict disease progression, explaining individual differences in the future trajectory of disease (i.e. prognostic from baseline) and the variation in cognitive decline over time (i.e. mediating mechanisms).

### Analysis

The analysis of MEG parameters will occur in two stages. Stage 1 analyses will identify baseline, disease-sensitive MEG parameters and assess their reliability using test-retest data. Stage 2 analyses will assess the sensitivity of these parameters to longitudinal change.

Stage 1 analyses will assess the sensitivity of the cross-sectional MEG parameters to group effects (using parametric or non-parametric frequentist tests and receiver operating characteristic analyses and their Bayesian analogues), their correlation to baseline cognition (using Pearson or Spearman’s rank correlation coefficient analyses) and their test-retest reliability (using intraclass correlation coefficient analyses). Stage 2 longitudinal analyses will assess how these neurophysiological markers change over ∼12 months (using linear mixed models of change) and compare their sensitivity (as accuracy and effects sizes) to disease progression with MRI and cognitive measures. Using multiple regression models, we will identify baseline biomarkers that are predictive of disease progression. *Post-hoc* exploratory analyses will examine the relationship between cognitive and imaging biomarkers as a function of: (i) variations in tau haplotype, (ii) polymorphisms of genes related to plasticity (e.g. brain-derived neurotrophic factor), (iii) Alzheimer’s disease risk alleles (e.g. apolipoprotein e4), (iv) polygenic risk scores of Alzheimer’s disease, and (v) plasma tau levels.

## METHODS

### Study Design

This is a repeated-measures, observational-design study with 130 participants tested at two sites: Cambridge University and Oxford University. The protocol includes two stages (Figure 1). The first, cross-sectional stage consists of baseline and test-retest sessions. Baseline sessions are comprised of clinical and neuropsychological assessments, an MRI scan and a MEG combined with EEG (M/EEG) scan. For the test-retest session, a subset of patients returns for a second M/EEG scan approximately two weeks after the first. Stage 2, at 12-months, repeats baseline sessions, followed by a 24-month clinical and neuropsychological reassessment of patients.

**Figure 1.**
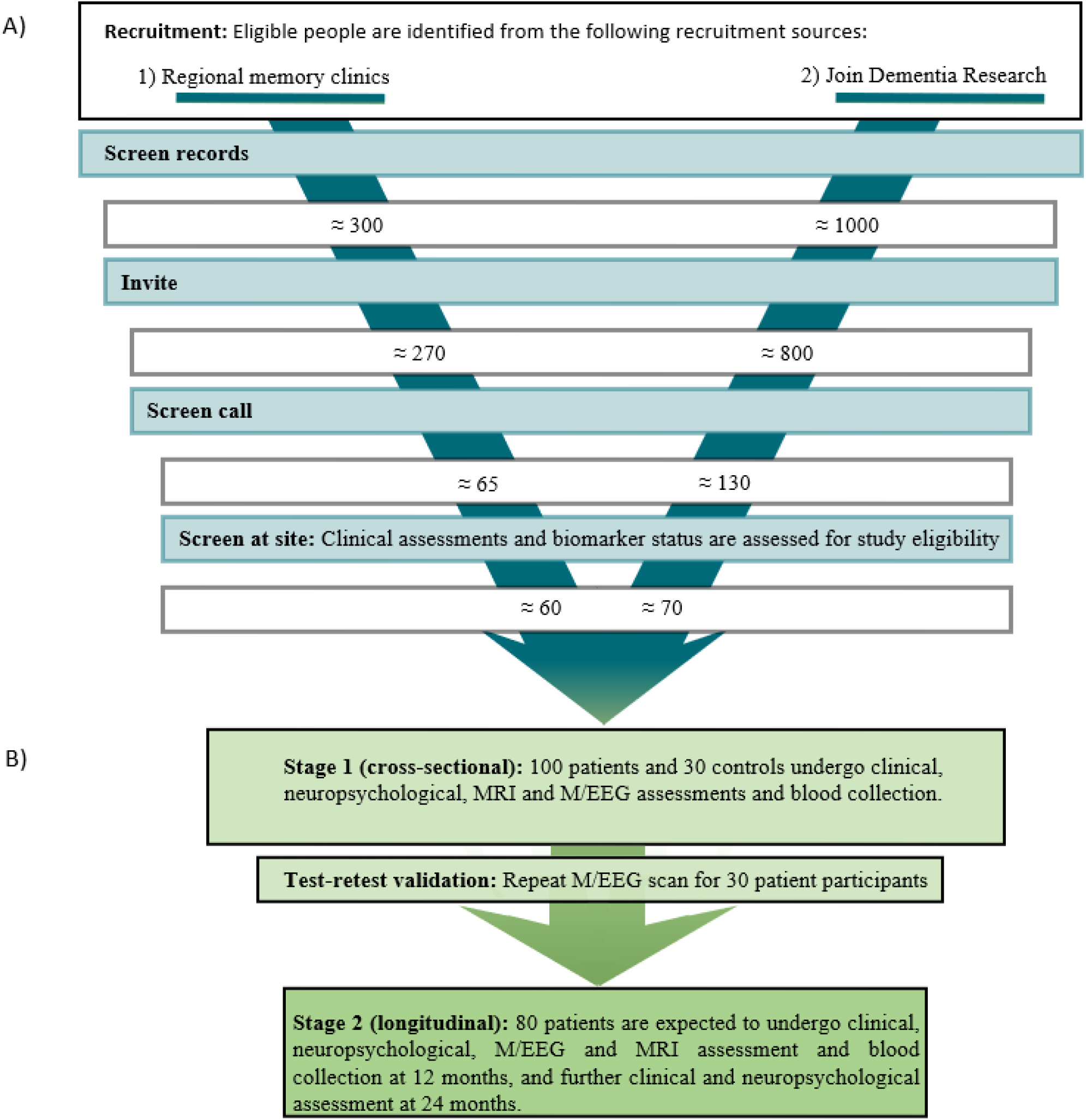
Participant flow chart. **A) Recruitment strategy**: Electronic screening of records from regional memory clinics and Join Dementia Research generates approximately 300 potential patient and 1000 potential patient and control participants who are invited to telephone screening. People who remain eligible are screened further on-site to identify 100 patient and 30 control participants. **B) Study stages**: Stage 1 consists of the cross-sectional baseline assessments and, for 30 patients, a repeat of the M/EEG assessment two weeks after the first. Only patients continue to the longitudinal stage 2 of the study, with repeat 12 and 24 months after baseline. Attrition of 20% is expected at stage 2. M/EEG=Magnetoencephalography combined with electroencephalography imaging, MRI=Magnetic resonance imaging

### Participant recruitment and selection

Participants will be aged between 50 and 85 years, with similar numbers of men and women, with symptomatic mild cognitive impairment or Alzheimer’s Disease (n=100) or normal cognition (n=30, Figure 1). Potential participants are identified using local registry data, regional memory clinics and Join Dementia Research. People completing other observational studies may also be invited to screening.

A participant information sheet detailing the study procedures is provided to candidate participants and a study partner (their ‘informant’). The informant is someone who regularly sees the participant and is willing to attend and complete the assessments. Candidate participants are invited to a screening appointment where any further questions they may have are answered before they provide written, informed consent. The screening appointment allows further assessment of eligibility according to inclusion and exclusion criteria (Table 1). Eligible people proceed to Stage 1.

**Table 1.**
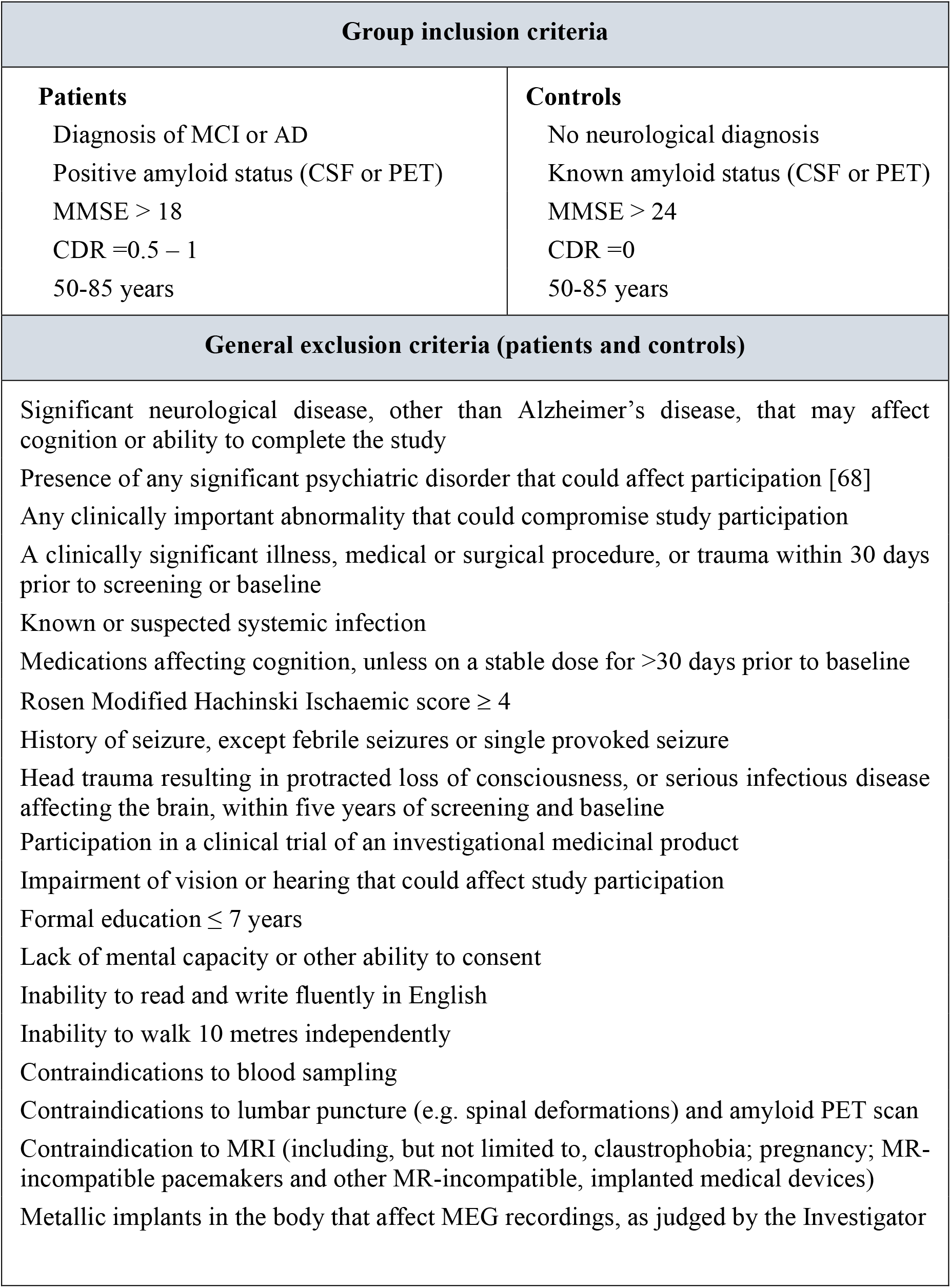
Group inclusion criteria and general exclusion criteria. MCI=Mild Cognitive Impairment, AD=Alzheimer’s Disease, MMSE=Mini Mental State Examination, CDR=Clinical dementia rating

| Group inclusion criteria |  |
| --- | --- |
| **Patients**<br>Diagnosis of MCI or AD<br>Positive amyloid status (CSF or PET)<br>MMSE > 18<br>CDR =0.5 – 1<br>50-85 years | **Controls**<br>No neurological diagnosis<br>Known amyloid status (CSF or PET)<br>MMSE > 24<br>CDR =0<br>50-85 years |
| General exclusion criteria (patients and controls) |  |
| Significant neurological disease, other than Alzheimer's disease, that may affect cognition or ability to complete the study<br>Presence of any significant psychiatric disorder that could affect participation [68]<br>Any clinically important abnormality that could compromise study participation<br>A clinically significant illness, medical or surgical procedure, or trauma within 30 days prior to screening or baseline<br>Known or suspected systemic infection<br>Medications affecting cognition, unless on a stable dose for >30 days prior to baseline<br>Rosen Modified Hachinski Ischaemic score $\geq 4$<br>History of seizure, except febrile seizures or single provoked seizure<br>Head trauma resulting in protracted loss of consciousness, or serious infectious disease affecting the brain, within five years of screening and baseline<br>Participation in a clinical trial of an investigational medicinal product<br>Impairment of vision or hearing that could affect study participation<br>Formal education $\leq 7$ years<br>Lack of mental capacity or other ability to consent<br>Inability to read and write fluently in English<br>Inability to walk 10 metres independently<br>Contraindications to blood sampling<br>Contraindications to lumbar puncture (e.g. spinal deformations) and amyloid PET scan<br>Contraindication to MRI (including, but not limited to, claustrophobia; pregnancy; MR-incompatible pacemakers and other MR-incompatible, implanted medical devices)<br>Metallic implants in the body that affect MEG recordings, as judged by the Investigator |  |

**Table 2.** Blood sample processing guidelines.

|  |  |
| --- | --- |
| **Plasma** | The sample in the sodium citrate tube is centrifuged at 2000g for 10 minutes at room temperature. The plasma is aliquoted into 1.4 ml matrix tubes and stored at -80°C. |
| **Serum** | At least 30 minutes after blood collection, the sample in the serum separator tube is centrifuged at 2000g for 10 min at room temperature. The serum is aliquoted into 1.4ml matrix tubes and stored at -80°C.<br>Similarly, the sample in the S-monovette tube is centrifuged at 2000g for 10 minutes at room temperature at least 30 minutes after collection. The sample is aliquoted into 2ml Sarstedt tubes. |
| **DNA** | The first EDTA tube is frozen as soon as possible and stored at -80°C for later DNA extraction and analysis. |
| **Plasma and buffy coat** | The second EDTA tube is centrifuged immediately at 2500g for 15 minutes at room temperature. The plasma and buffy coat are aliquoted into separate 1.4 ml matrix tubes and stored at -80°C. |
| **RNA** | The Paxgene tubes are stored at room temperature for 2 hours before being frozen upright at -20°C for 24 hours and then stored at -80°C. |

### Sample Size and Power

With power of 80% and α=0.05, a one-tailed test of the cross-sectional NTAD data should detect (i) medium, group-wise effect sizes of >0.5 and (ii) correlations with disease severity of >0.25. With 20% attrition in follow up and α=0.05, within-sample effect size >0.28 achieves power 80% (Figure 2).[34]

**Figure 2.**
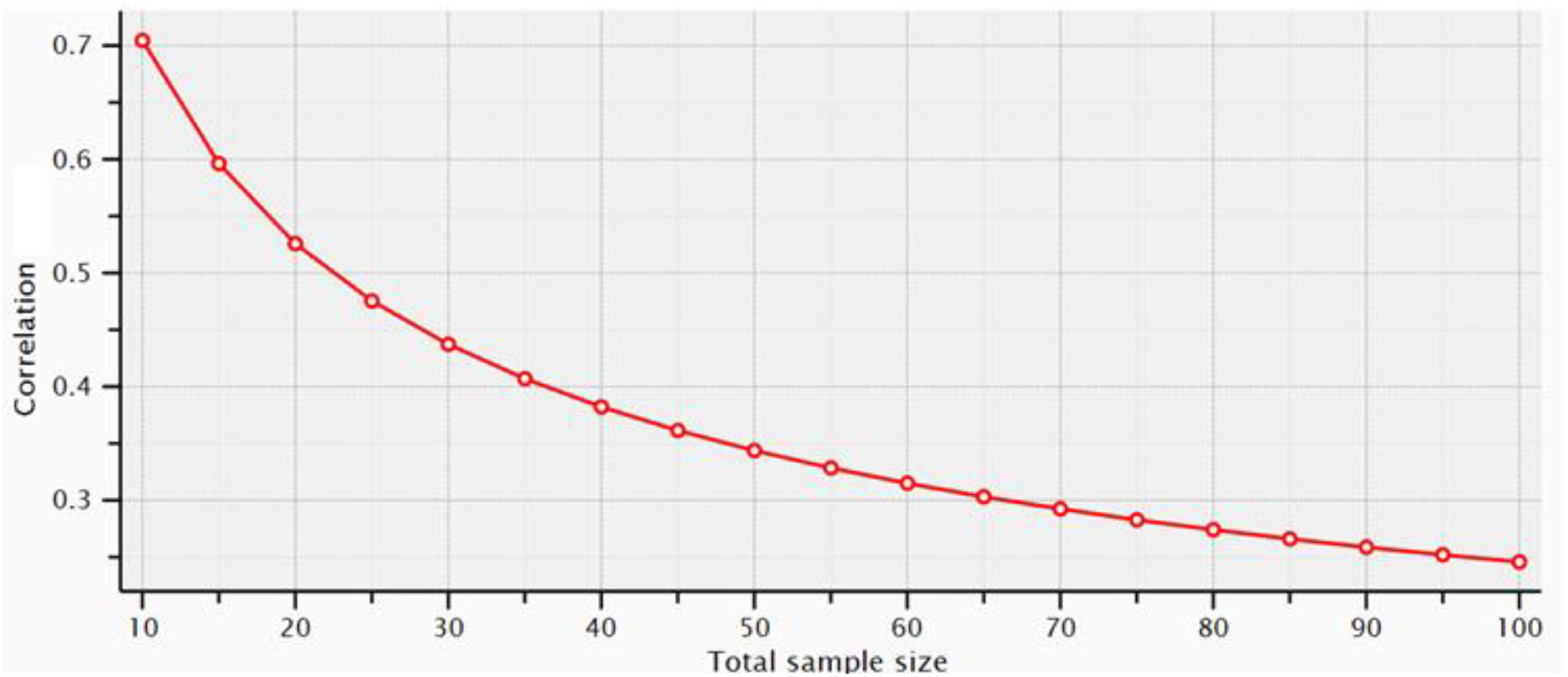
Effect sizes required to detect longitudinal correlations with disease severity at different attrition rates. (resulting in reduced longitudinal sample size) with 80% power and α=0.05. We expect <20% attrition.

## STUDY PROCEDURES

### Overview of protocol stages

#### Screening

Potential participants are identified by electronically screening registry data from Join Dementia Research and regional memory clinics. Prospective participants are screened further on-site. After written, informed consent, a doctor administers the clinical interview and Haschinski ischemic scale. The remaining clinical assessments are completed by a member of the research team and include physiology and blood sampling. If amyloid status is unknown, participants proceed to either cerebrospinal fluid examination or amyloid PET imaging according to participant preference and eligibility. For patients, where participants’ amyloid status has been confirmed at any timepoint previously, a positive result enables participation, a negative result excludes participation.

#### Stage 1: Baseline

One hundred people with mild cognitive impairment or Alzheimer’s disease and 30 neurologically normal people proceed to baseline assessment. Participants undergo structured neuropsychological assessment, an M/EEG scan and MRI imaging of brain structure and function over two sessions (or three by preference).

#### Stage 1: Two-week M/EEG retest

We invite 30 people from the patient group (i.e. mild cognitive impairment or Alzheimer’s disease) to repeat the M/EEG scan between two and four weeks after the first scan.

Invitations are prioritised to people who can most readily attend the additional session (e.g. considering distance) until the target sample size is reached.

#### Stage 2: Annual follow-up 1

Participants in the patient group repeat the clinical and neuropsychological assessments, M/EEG scan, MRI scan and blood collection at 12 months after baseline.

#### Stage 2: Annual follow-up 2

Clinical and neuropsychological assessments are repeated at 24 months for participants in the patient group.

The timeline for the study is illustrated in Figure 3.

**Figure 3.**
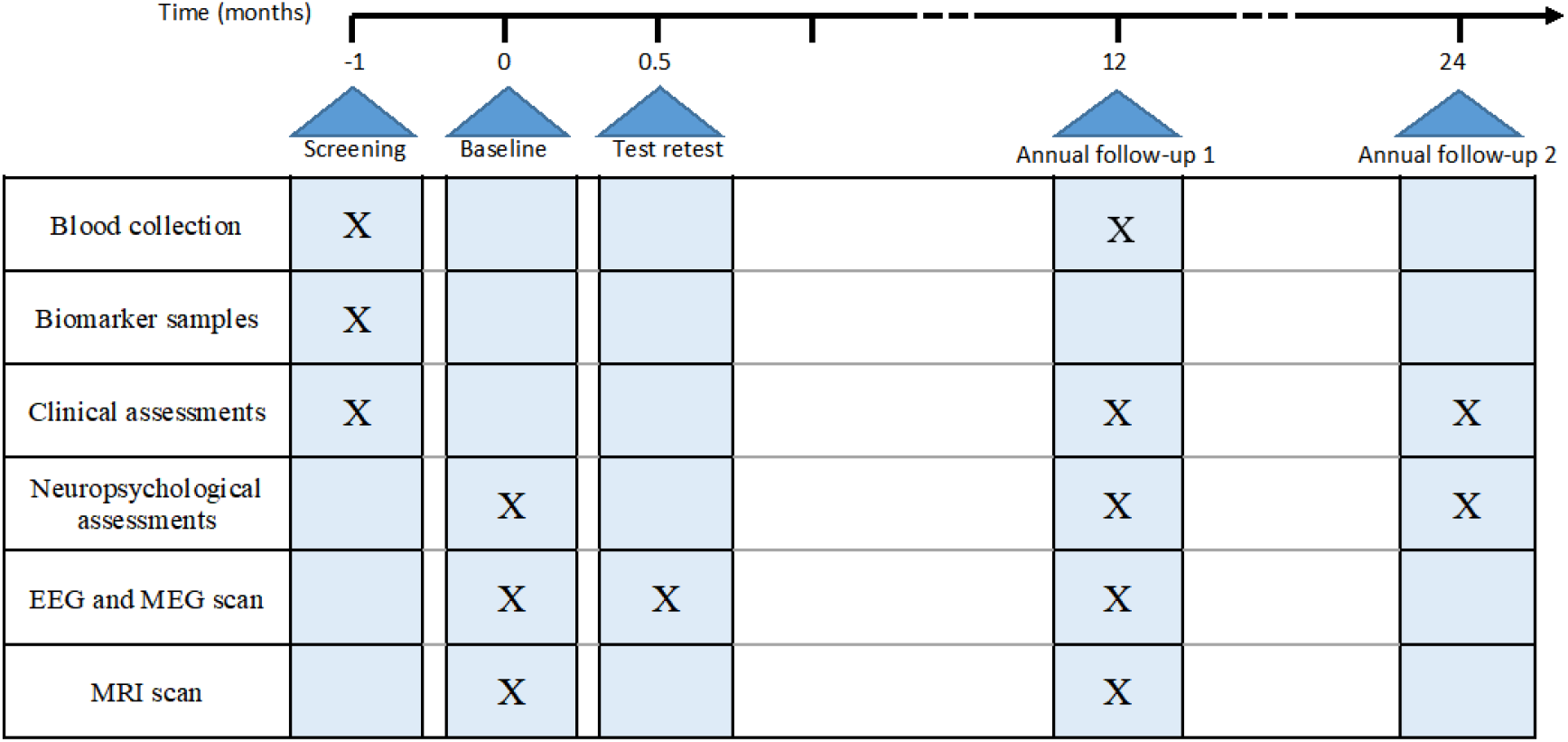
Timeline summary of study procedures.

### Blood Samples

Participants are asked to consume only water for two hours prior to blood collection. At baseline and follow-up, blood is drawn in the following order: 2.7ml in Sodium Citrate Tubes (plasma), 5ml in Serum Separator Tubes (serum), 10ml in an EDTA tube (DNA), 10ml in an EDTA tube (plasma and buffy coat) and 2.5ml in each of 2 PAXgene tubes (RNA). Once filled, the EDTA and PAXgene tubes are gently inverted 10 times. From April 2021, 4.9ml of blood is collected for SARS-CoV-2 serology. The samples are processed and frozen according to the guidelines for the constituents of interest in

### Biomarkers for Alzheimer disease pathology

Clinical criteria are insufficient to reliably diagnose the presence of Alzheimer’s disease pathology.[35,36] We seek additional biomarker evidence of the presence of Alzheimer’s pathology, using either cerebrospinal fluid or PET imaging according to participant preference and eligibility.[37]

#### Cerebrospinal fluid

Cerebrospinal fluid is obtained by lumbar puncture and collected in polypropylene tubes. Within one hour of collection, the cerebrospinal fluid is centrifuged, separated and the supernatant frozen to −80°C for later batched analysis using a chemiluminescent enzyme immunoassay for total tau, phosphorylated tau and amyloid beta 1-42 levels. Positive amyloid status is indicated by a total tau to amyloid beta 1-42 ratio >1, and amyloid beta 1-42 concentration <450pg/ml.

#### Amyloid PET

Participants receive a 300 MBq bolus injection of florbetaben and are scanned 80-100 minutes post-injection on a GE Signa PET/MR scanner at the Wolfson Brain Imaging Centre, Cambridge or a GE D710 PET/CT scanner at the Churchill Hospital, Oxford. The centiloid method is used to classify the florbetaben scans as amyloid positive with centiloid >1.19.[38,39]

### Clinical Assessments

Clinical assessments are completed at a clinical research centre in one or two sessions.

#### Clinical Interview

A study clinician interviews the participant and study partner. The clinician follows a structured interview that covers socio-demographic factors, including age and years of education; lifestyle factors; family history of dementia; medical history, including information on significant medical conditions with date of onset; and concomitant medication usage, with details about dosage and duration.

#### Addenbrooke’s Cognitive Examination

The revised Addenbrooke’s Cognitive Examination (ACE-R) evaluates orientation, memory, verbal fluency, language and visuo-spatial domains.[40] It is administered and scored according to the ACE-R Administration and Scoring Guide (2006). Alternate versions at each visit reduce practice effects.

#### Mini-Mental State Examination

The Mini-Mental State Examination was acquired.[41]

#### Clinical Dementia Rating

The Clinical Dementia Rating quantifies the severity of dementia through a structured interview.[42–44] The interviewer rates participant impairments in 6 categories: memory, orientation, judgment and problem solving, community affairs, home and hobbies and personal care. The global CDR is derived from these ratings.[43]

#### Haschinski Ischaemic Score

The Rosen modification of Haschinski’s Ischaemic Score seeks to differentiate between primary progressive dementias, including Alzheimer’s disease and multi-infarct dementia.[45] A study clinician uses information from medical history, physical and neurological examination and medical records to determine the score. Scores below 4 indicate a low likelihood of vascular disease as the cause of dementia.

#### Self-reported questionnaires

The participant completes the 30-item Geriatric Depression Scale,[46] 40-item Spielberger State-Trait Anxiety Inventory,[47] and 11-item Pittsburgh Sleep Quality Index.[48] The study partner completes the 30-item Amsterdam Instrumental Activity of Daily Living Questionnaire (short version)[49] and Mild Behavioral Impairment Checklist.[50]

#### Physiological measures

Height, weight, hip-waist ratio, blood pressure, pulse rate and temperature are measured for each participant using a stadiometer, electronic weight scales and stretch-resistant tape.

### Neuropsychological Assessment

The neuropsychological test battery closely resembles the Deep and Frequent Phenotyping study[51] and IMI-European Prevention of Alzheimer’s Disease,[52,53] including the Pre-Alzheimer Cognitive Composite.[54] The assessments take place in a private, testing room in one session with breaks as needed (see Table 3).

**Table 3.** Order of neuropsychological assessments. RBANS=Repeatable Battery for the Assessment of Neuropsychological Status, CANTAB = Cambridge Neuropsychological Test Automated Battery

| Order of assessments | Duration (minutes) |
| --- | --- |
| 1. List learning (RBANS subtest)<br>2. Story memory (RBANS subtest)<br>3. Figure Copy (RBANS subtest)<br>4. Line orientation (RBANS subtest)<br>5. Picture naming (RBANS subtest)<br>6. Semantic fluency (RBANS subtest)<br>7. Digit span forwards (RBANS subtest)<br>8. Digit span backwards<br>9. Coding (RBANS subtest)<br>10. List recall (RBANS subtest)<br>11. List recognition (RBANS subtest)<br>12. Story recall (RBANS subtest)<br>13. Figure recall (RBANS subtest) | ≈ 40 |
| *Break* | ≈ 15 |
| 14. Free and cued selective reminding test<br>15. Logical memory part 1<br>16. National adult reading test<br>17. Digit symbol substitution<br>18. Trails B<br>19. Logical memory part 2 | ≈ 45 |
| *Break* | ≈ 15 |
| 20. Reaction time task (CANTAB subtest)<br>21. Paired associates learning (CANTAB subtest)<br>22. Rapid visual processing (CANTAB subtest)<br>23. Spatial working memory (CANTAB subtest)<br>24. Four mountains task | ≈ 30 |

### Repeatable Battery for the Assessment of Neuropsychological Status

Repeatable Battery for the Assessment of Neuropsychological Status (RBANS) measures cognitive decline in 5 domains: immediate memory, visuospatial, language, attention and delayed memory.[55] Participants receive alternate forms at repeated assessments. The immediate memory index comprises the *list learning subtest*, with immediate recall of 10-items over 4 trials and the *story memory subtest*, with immediate recall of a 12-item story over two trials. The visuospatial index comprises the *figure copy subtest*, which involves copying a geometric figure and the 10-item, *line orientation subtest*. The language index comprises the 10-item, *picture naming subtest* and the *semantic fluency subtest*, where the participant must name as many exemplars of the given semantic category as they can in 60 seconds. The attention index comprises the *digit span forwards subtest*, involving immediate repetition of increasing digit strings and the *coding subtest*, scored as the total number of correctly coded numbers generated using an item-to-number code within 90 seconds. The delayed memory index comprises the *list recall subtest*, involving free recall of the list learning task; *list recognition*, where the participant decides whether a word was included in the list learning task; *story recall*, where the participant freely recalls the story memory task; and *figure recall*, where the participant draws from memory the figure presented in the figure copy task.

#### Digit Span Backwards

Digit span backwards, from the Wechsler adult intelligent scale,[56] is used in conjunction with RBANS digit span forwards. It follows the RBANS digit span format except that the participant is asked to immediately repeat in reverse order.

#### Free and Cued Selective Reminding Test

The Free and Cued Selective Reminding Testassesses episodic memory and distinguishes retrieval from storage deficits.[57] Sixteen pictured items are encoded during the initial learning phase, where the participant identifies and names items responding to unique semantic cues. After a short delay, the participant freely recalls all items. The interviewer prompts for each item not recalled using the unique semantic cues from the learning phase. Participants receive alternate forms at repeated assessments. This test forms part of the Pre-Alzheimer Cognitive Composite.

#### Logical Memory

Logical Memory, taken from the third edition of the Wechsler memory scale, assesses episodic memory.[58] The participant immediately recalls short stories they have been read. After a 30-minute delay with intervening tests, the participant freely recalls the stories and answers yes or no questions that test story recognition. This test forms part of the Pre-Alzheimer Cognitive Composite.

#### National Adult Reading Test

The national adult reading test (second edition) estimates premorbid intelligence,[59] from printed irregular words.

#### Digit Symbol Substitution

The digit symbol substitution from the Wechsler Adult Intelligence Scale assesses processing speed and attention.[56] The participant has 90 seconds to code as many correct symbols as possible corresponding to presented numbers by using the given number-to-symbol code. This test forms part of the Pre-Alzheimer Cognitive Composite.

#### Trails Making Test B

The Trails Making Test B assesses executive function, attention and processing speed.[60] Following a practice sample to ensure the participant understands the task, the participant is presented with the test comprising 25 encircled numbers which they must connect by alternating between numbers and letters. The time it takes to complete the sample and any errors made are recorded.

#### Cambridge Neuropsychological Test Automated Battery

The tablet-based Cambridge Neuropsychological Test Automated Battery test battery assesses processing speed, episodic memory, attention, working memory and executive function.[61] The reaction time task (simple and five choice variant) assesses processing speed. The participant holds down a response button and must release this to respond to a target on screen. The paired associates learning task (standard variant) assesses episodic memory. The participant learns associations between patterns and their locations. There is an initial learning stage followed by immediate recall. The rapid visual processing task (3 target variant) assesses attention. Single digits appear on the screen and the participant responds when they see a string matching the target sequences. The spatial working memory task (standard variant) assesses working memory and executive function. The participant searches inside multiple boxes on the screen to find and collect tokens. Tokens do not appear in the same box twice.

#### Four Mountains Task

The tablet-based four mountains task assesses allocentric spatial processing.[62] During a learning phase of each trial, the participant learns the topographical layout of 4 mountains presented in a computer-generated landscape. Following a delay, the participant is presented with four alternative images and must identify the target image that matches the topographical mountain layout of the image presented in the learning phase of the trial, but with potentially altered colours, textures and points of view.

### Neurophysiology (M/EEG)

M/EEG data are collected simultaneously in a magnetically-shielded room. At Cambridge, data are collected using the Elekta VectorView system from 2017 to December 2019, with 204 planar gradiometers and 102 magnetometers and a 70-channel Easycap. Stage 1 scans and the first 12 follow-up scans used the same scanner. The MEGIN Triux Neo M/EEG scanner is used from March 2020 onwards with the same sensor configuration as the VectorView and a 64-channel Easycap. At Oxford, the MEGIN Triux Neo M/EEG scanner and an EasyCap 60 channel BrainCap for MEG with an augmented 10/20 layout are used for all data collection. M/EEG data are collected at 1000Hz.

The position of the standard fiducial points, >300 additional head points, five head position indicator coils and the EEG electrodes are recorded using the Polhemus digitisation system. The head position indicator coils measure head position within the MEG helmet. Three pairs of bipolar electrodes record electrocardiogram data, with electrodes placed on the right clavicle and left lower rib, and vertical and horizontal electrooculogram data, with electrodes placed above the left eyebrow, below the left eye and lateral canthus of each eye. A reference electrode is placed on the left side of the nose and a ground electrode is placed on the left cheek. During the seated scan, the participant rests or performs simple task using a button box to respond. The participant wears non-magnetic earphones with sound delivered though plastic tubes and, if necessary, non-magnetic glasses. Prior to M/EEG a Snellen eye test and pure tone audiometry assess sight and hearing thresholds. The tasks are carried out in the following order for each M/EEG session. The study will identify a harmonised MEG/EEG pre-processing pipeline, to be confirmed before baseline data acquisition is completed. Simple audio-visual task

The participant fixates on a red central fixation dot and responds each time an auditory or visual stimulus is presented (see Figure 4a). Auditory tones (n=100) of 300, 600 or 1200Hz frequency are presented for 300-ms duration after a blank interval of 1000ms. Visual stimuli are concentric black and white circles that appear for 300ms after 3000ms. Participants respond as quickly as possible after detecting a stimulus. Filler trials (n=30) containing only the red fixation dot are also included. Visual and auditory trials are randomly interspersed. Ten initial practice trials are also included to familiarise participants to the task.

**Figure 4.**
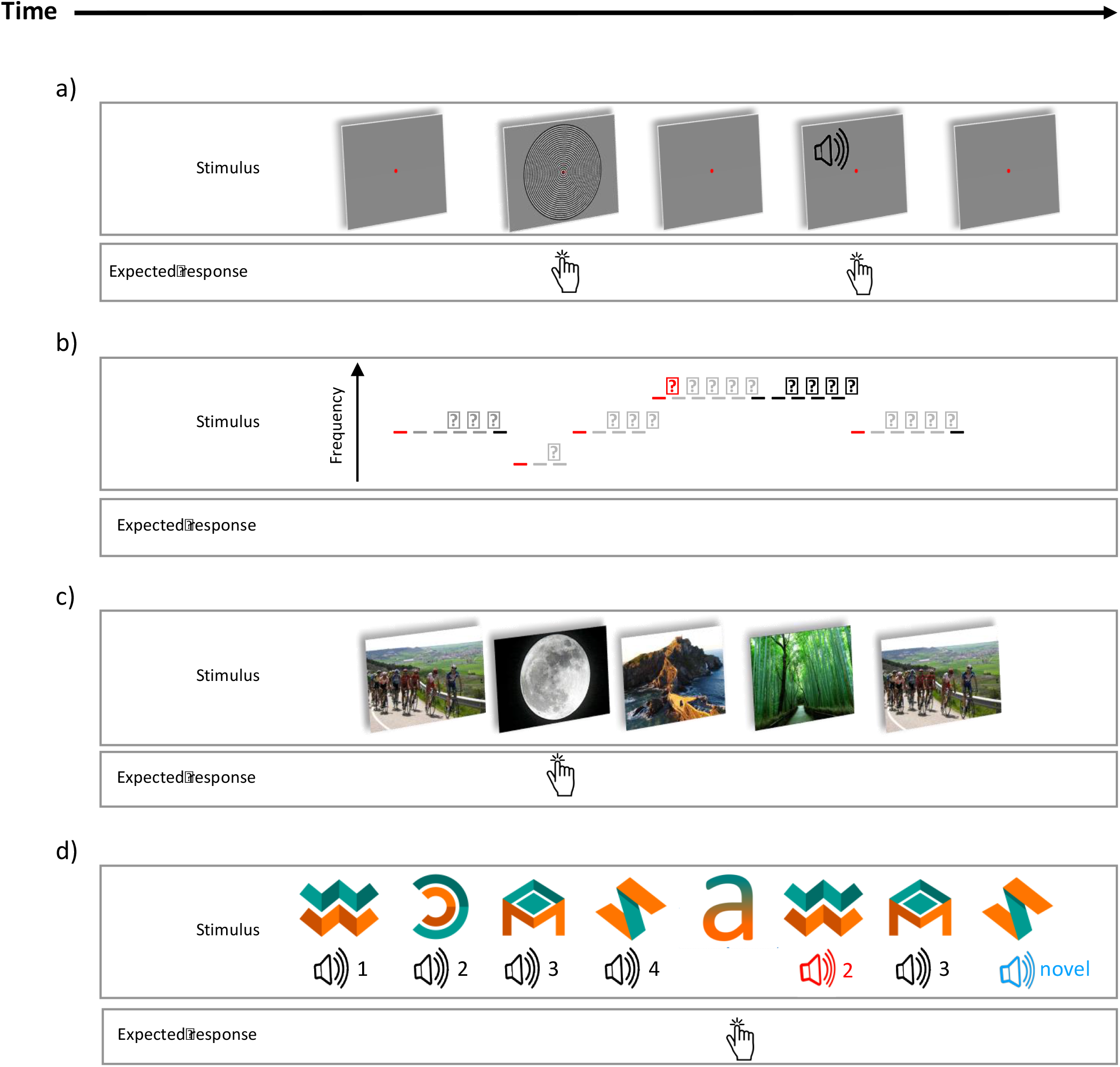
**a) Audio-visual task:** Participants visually fixate on a red dot and press a button whenever they hear or see something. **(b) Mismatch-negativity task:** Participants passively watch a nature documentary while listening to tones through the earpieces. Red dashes represent deviant tones and black dashes represent standard tones (from the sixth repetition). **(c) Scene repetition task:** Participants view scenes and press the button when they see a scene containing a moon. Each non-target scene repeats once during the task. **(d) Cross-modal oddball task:** Participants view object-tone pairs consisting of 4 learnt (standard) pairs (the standard sounds are black in the figure); associative-deviant, object-tone pairs (the associative-deviant sound is coloured red in the figure); and standard objects presented with novel tones (the novel-deviant tones are blue in the figure). The participant presses a button when they see the letter ‘a’.

#### Auditory Mismatch-negativity Task

The roving auditory mismatch negativity task is designed to elicit error responses to novel, deviant tones followed by rapid plasticity as predictions are updated upon repetition of the deviant stimulus.[63,64] The participant passively watches a muted nature documentary. Through earpieces, they hear binaural, in-phase sinusoidal tones >60dB above the average auditory threshold with a duration of 100ms and stimulus onset asynchrony of 500ms. The frequency of each tone is the same within a block but different between the blocks. Blocks range from 400 to 800 Hz. The number of tones per blocks varies from 3 to 11, according to a truncated exponential distribution. The first tone of each block is called a deviant tone and the sixth repetition is called a standard tone (see Figure 4b).

#### Scene Repetition Task

In this passive memory test, participants view a series of complex scenes (landscapes and cityscapes) and press a button only when they see a scene containing a moon (n=26, different moon images). Target scenes containing a moon require participants to attend to every scene but are not of interest; the main interest is the difference between initial and repeated presentation of the non-target scenes. Each scene is presented for 800ms and preceded by a fixation cross presented for 200ms on average (100-300ms). Scenes are pseudo-randomly intermixed with the constraint that there are 10 initial ‘burn-in’ complex scenes with 2 moon target scenes, followed by 256 complex scenes presented twice with 14-93 (median=42) intervening scenes between the first and repeat presentation (see Figure 4c).

#### Cross-modal Oddball Task

The task assesses hippocampal-dependent paired associates learning.[65,66] The trials comprise a visual abstract “object” for 700ms and a sound presented for 400ms, starting 300ms after trial onset. The inter-stimulus interval averages 300ms. During the initial training period (80 trials), participants learn the association between four standard pairs of visual objects and sounds. The main task consists of 770 bimodal trials and 40 unimodal, target trials. The trials are randomly intermixed. The bimodal trials consist of: the standard, learnt pairs of objects and sounds (n=670); standard objects paired with a novel sound (n=50); and mismatched object-sound pairs, where the sound from a pre-learnt, standard pair is presented with an object from a different learnt, standard pair (n=50). For unimodal trials, the participant presses a button when they see a fifth visual object (the letter ‘a’, see Figure 4d) which ensures the task is attended. The task is followed by a 10-item assessment in which a standard sound is presented and the participant reports which of four presented objects the sound was paired with most often during the task.

#### Eyes-open Resting State

The participant is presented with a small central fixation cross and is given the following instructions: ‘In the next 5 minutes, we will do a recording as you rest. Please clear your mind, relax, and try not to think of anything in particular. Please stay awake and focus your eyes on the cross at the centre of the screen’.

#### Eyes-closed Resting State

The participant is given the following instructions: ‘In the next 5 min, we will do a recording as you rest. Please clear your mind, relax, and try not to think of anything in particular. Please close your eyes but do not fall asleep.’

### Magnetic Resonance Imaging

The MRI sequences use 3T Siemens PRISMA scanners, at the MRC Cognition and Brain Science Unit, Cambridge, and the Oxford Centre for Human Brain Activity, Oxford. The sequences in order of acquisition are: localiser, T1-weighted, T2 Fluid Attenuated Inversion Recovery (FLAIR), T2*-weighted, T2-weighted with fat saturation, diffusion weighted imaging, quantitative susceptibility mapping, resting state eyes open, hippocampal subfields, and 3D arterial spin labelling. Details of the sequences are in Table 4.

**Table 4.**
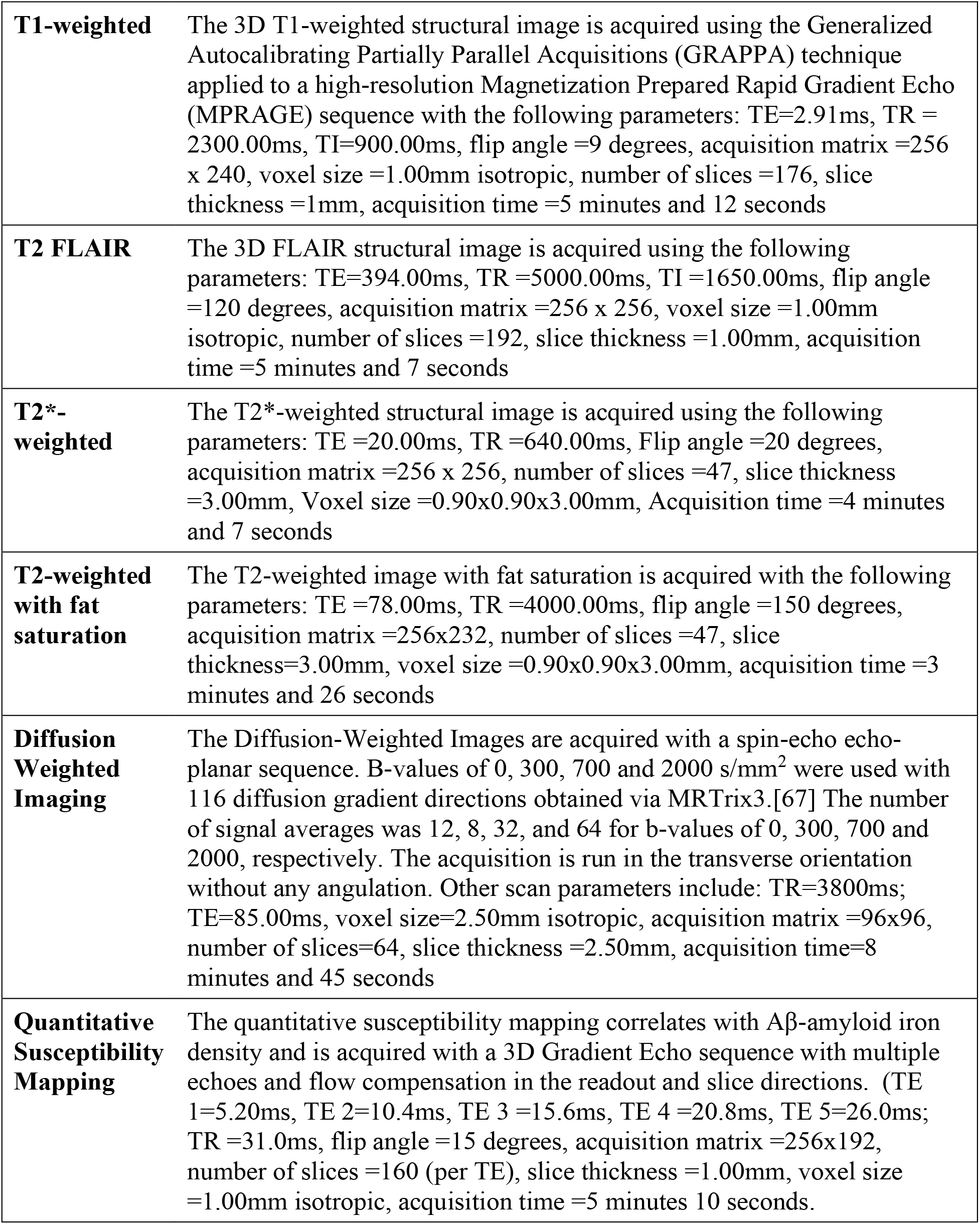
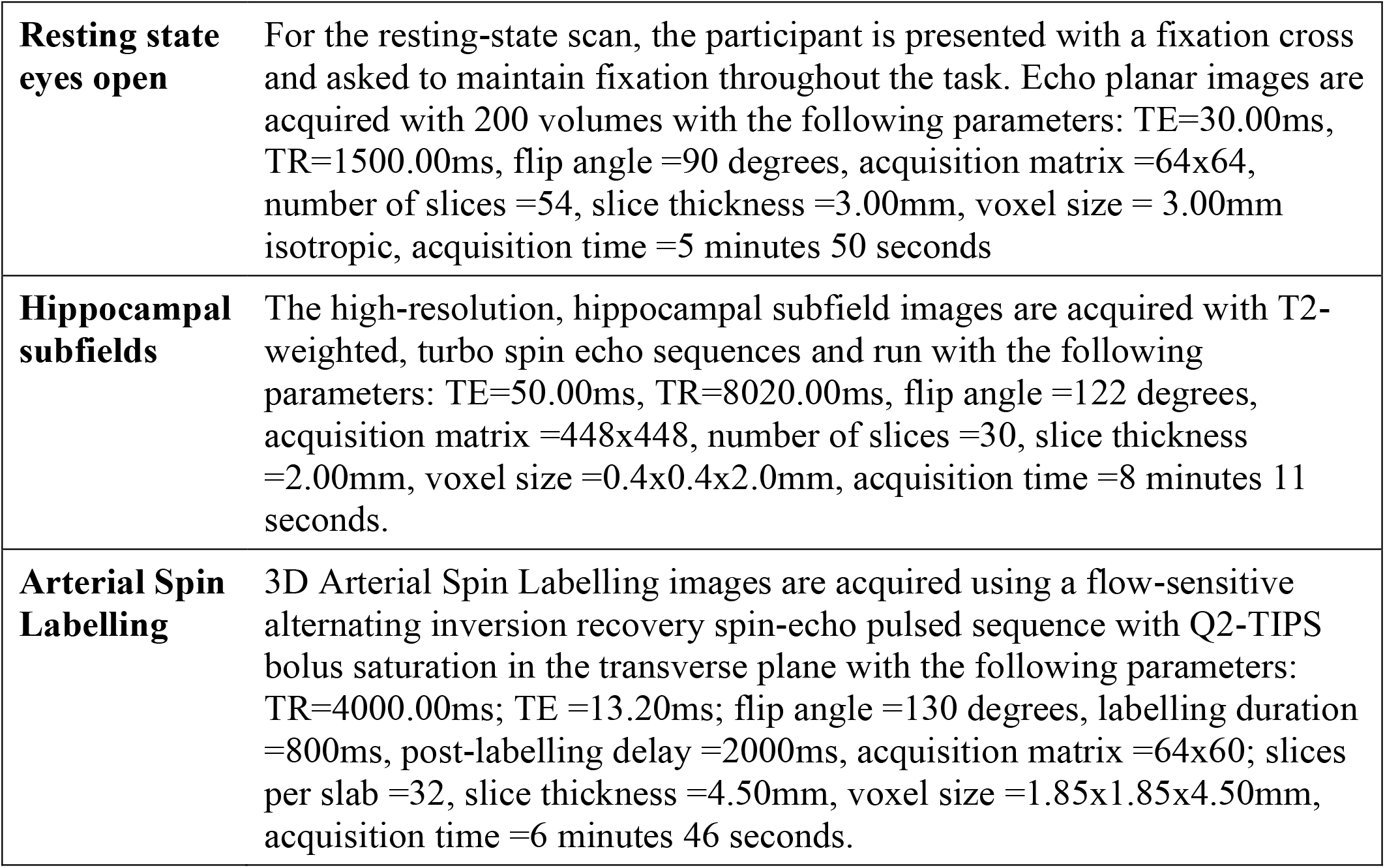
MRI sequence parameters. TE=Echo time, TR=Repetition time, TI=Inversion time

## CURRENT STATUS

The study is currently active at both sites.

## ETHICS AND DISSEMINATION PLAN

The study has received a favourable opinion from the East of England – Cambridge Central Research Ethics Committee (REC reference 18/EE/0042). Imaging data and clinical severity scores are hosted by the Dementias Platform UK Imaging Platform (https://portal.dementiasplatform.uk), using XNAT technology (https://www.xnat.org). The data will be made available by a managed access process through the DPUK, subject to constraints to preserve confidentiality and privacy.

## CONCLUSION

With a pressing need to evaluate new treatments for Alzheimer’s disease, measures that are sensitive to changes in the underlying neurophysiology of the human disease are needed. The NTAD study establishes a dataset that will allow detailed assessment of neurophysiological measures of annual progression, linked to cognitive and MRI changes and baseline blood and DNA. The unique design of the NTAD study enables an assessment of MEG as a potential biomarker in Alzheimer’s disease intervention trials.

## Data Availability

Imaging data and clinical severity scores are hosted by the Dementias Platform UK Imaging Platform (https://portal.dementiasplatform.uk), using XNAT technology (https://www.xnat.org). The data will be made available by a managed access process through the DPUK, subject to constraints to preserve confidentiality and privacy.

## FOOTNOTES

### Authors’ contributions

JHL and JBR drafted the manuscript, with review and contribution from all authors. JBR is the chief investigator of the study. RNH, ACN, MW, KDS and VR are principal investigators, contributing to conception and design. EK led protocol development and governance. JBR, MW, ACN, RNH, JI, SL, MP and KDS conceived and designed the study.

## REFERENCES

1 LaFerla FM, Green KN. Animal models of Alzheimer disease. Cold Spring Harb Perspect Med 2012;2:1–13. doi:10.1101/cshperspect.a006320

2 Drummond E, Wisniewski T. Alzheimer’s disease: experimental models and reality. Acta Neuropathol. 2017;133:155–75. doi:10.1007/s00401-016-1662-x

3 Cummings JL, Morstorf T, Zhong K. Alzheimer’s disease drug-development pipeline: Few candidates, frequent failures. Alzheimer’s Res Ther 2014;6:1–7. doi:10.1186/alzrt269

4 Yiannopoulou KG, Anastasiou AI, Zachariou V, et al. Reasons for failed trials of disease-modifying treatments for alzheimer disease and their contribution in recent research. Biomedicines. 2019;7. doi:10.3390/biomedicines7040097

5 Anderson RM, Hadjichrysanthou C, Evans S, et al. Why do so many clinical trials of therapies for Alzheimer’s disease fail? Lancet. 2017;390:2327–9. doi:10.1016/S0140-6736(17)32399-1

6 Mehta D, Jackson R, Paul G, et al. Why do trials for Alzheimer’s disease drugs keep failing? Expert Opin Investig Drugs 2017;26:735–9. doi:10.1080/13543784.2017.1323868.Why

7 Frisoni GB, Fox NC, Jack CR, et al. The clinical use of structural MRI in Alzheimer disease. Nat Rev Neurol 2010;6:67–77. doi:10.1038/nrneurol.2009.215

8 Holland D, McEvoy LK, Dale AM. Unbiased comparison of sample size estimates from longitudinal structural measures in ADNI. Hum Brain Mapp 2012;33:2586–602. doi:10.1002/hbm.21386

9 Leung KK, Barnes J, Ridgway GR, et al. Automated cross-sectional and longitudinal hippocampal volume measurement in mild cognitive impairment and Alzheimer’s disease. Neuroimage 2010;51:1345–59. doi:10.1016/j.neuroimage.2010.03.018

10 Cope TE, Rittman T, Borchert RJ, et al. Tau burden and the functional connectome in Alzheimer’s disease and progressive supranuclear palsy. Brain 2018;141:550–67. doi:10.1093/brain/awx347

11 Passamonti L, Rodríguez PV, Hong YT, et al. PK11195 binding in Alzheimer disease and progressive supranuclear palsy. Neurology 2018;90:e1989–96. doi:10.1212/WNL.0000000000005610

12 Passamonti L, Rodríguez PV, Hong YT, et al. 18F-AV-1451 positron emission tomography in Alzheimer’s disease and progressive supranuclear palsy. Brain 2017;140:781–91. doi:10.1093/brain/aww340

13 Leuzy A, Chiotis K, Lemoine L, et al. Tau PET imaging in neurodegenerative tauopathies—still a challenge. Mol. Psychiatry. 2019;24:1112–34. doi:10.1038/s41380-018-0342-8

14 Ossenkoppele R, Schonhaut DR, Schö M, et al. Tau PET patterns mirror clinical and neuroanatomical variability in Alzheimer’s disease. Brain 2016;139:1551–67. doi:10.1093/brain/aww041

15 Cai L, Innis R, Pike V. Radioligand Development for PET Imaging of beta-Amyloid (AD)-Current Status. Curr Med Chem 2006;14:19–52. doi:10.2174/092986707779313471

16 Bevan-Jones WR, Surendranathan A, Passamonti L, et al. Neuroimaging of inflammation in memory and related other disorders (NIMROD) study protocol: A deep phenotyping cohort study of the role of brain inflammation in dementia, depression and other neurological illnesses. BMJ Open 2017;7. doi:10.1136/bmjopen-2016-013187

17 Koffie RM, Hyman BT, Spires-Jones TL. Alzheimer’s disease: Synapses gone cold. Mol. Neurodegener. 2011;6:1–9. doi:10.1186/1750-1326-6-63

18 Palop JJ, Mucke L. Amyloid-Β-induced neuronal dysfunction in Alzheimer’s disease: From synapses toward neural networks. Nat. Neurosci. 2010;13:812–8. doi:10.1038/nn.2583

19 Kashyap G, Bapat D, Das D, et al. Synapse loss and progress of Alzheimer’s disease - A network model. Sci Rep 2019;9:1–9. doi:10.1038/s41598-019-43076-y

20 De Wilde MC, Overk CR, Sijben JW, et al. Meta-analysis of synaptic pathology in Alzheimer’s disease reveals selective molecular vesicular machinery vulnerability. Alzheimer’s Dement 2016;12:633–44. doi:10.1016/j.jalz.2015.12.005

21 Henstridge CM, Sideris DI, Carroll E, et al. Synapse loss in the prefrontal cortex is associated with cognitive decline in amyotrophic lateral sclerosis. Acta Neuropathol 2018;135:213–26. doi:10.1007/s00401-017-1797-4

22 Menkes-Caspi N, Yamin HG, Kellner V, et al. Pathological tau disrupts ongoing network activity. Neuron 2015;85:959–66. doi:10.1016/j.neuron.2015.01.025

23 Pinotsis DA, Schwarzkopf DS, Litvak V, et al. Dynamic causal modelling of lateral interactions in the visual cortex. Neuroimage 2013;66:563–76. doi:10.1016/j.neuroimage.2012.10.078

24 Adams NE, Hughes LE, Phillips HN, et al. GABA-ergic dynamics in human frontotemporal networks confirmed by pharmaco-magnetoencephalography. J Neurosci 2020;:1689–19. doi:10.1523/jneurosci.1689-19.2019

25 Sami S, Williams N, Hughes LE, et al. Neurophysiological signatures of Alzheimer’s disease and frontotemporal lobar degeneration: pathology versus phenotype. Brain 2018;141:2500–10. doi:10.1093/brain/awy180

26 Sitnikova TA, Hughes JW, Ahlfors SP, et al. Short timescale abnormalities in the states of spontaneous synchrony in the functional neural networks in Alzheimer’s disease. NeuroImage Clin 2018;20:128–52. doi:10.1016/j.nicl.2018.05.028

27 Puttaert D, Coquelet N, Wens V, et al. Alterations in resting-state network dynamics along the Alzheimer’s disease continuum. Sci Rep 2020;10:1–13. doi:10.1038/s41598-020-76201-3

28 Hughes LE, Henson RN, Pereda E, et al. Biomagnetic biomarkers for dementia: A pilot multicentre study with a recommended methodological framework for magnetoencephalography. Alzheimer’s Dement Diagnosis, Assess Dis Monit 2019;11:450–62. doi:10.1016/j.dadm.2019.04.009

29 Hughes LE, Nestor PJ, Hodges JR, et al. Magnetoencephalography of frontotemporal dementia: Spatiotemporally localized changes during semantic decisions. Brain 2011;134:2513–22. doi:10.1093/brain/awr196

30 Hughes LE, Rittman T, Robbins TW, et al. Reorganization of cortical oscillatory dynamics underlying disinhibition in frontotemporal dementia. Brain 2018;141:2486– 99. doi:10.1093/brain/awy176

31 Hughes LE, Rittman T, Regenthal R, et al. Improving response inhibition systems in frontotemporal dementia with citalopram. Brain 2015;138:1961–75. doi:10.1093/brain/awv133

32 Hughes LE, Rowe JB. The impact of neurodegeneration on network connectivity: A study of change detection in frontotemporal dementia. J Cogn Neurosci 2013;25:802– 13. doi:10.1162/jocn_a_00356

33 van Dellen E, de Waal H, van der Flier WM, et al. Loss of EEG Network Efficiency Is Related to Cognitive Impairment in Dementia With Lewy Bodies. Mov Disord 2015;30:1785–93. doi:10.1002/mds.26309

34 Erdfelder E, FAul F, Buchner A, et al. Statistical power analyses using G*Power 3.1: Tests for correlation and regression analyses. Behav Res Methods 2009;41:1149–60. doi:10.3758/BRM.41.4.1149

35 Ossenkoppele R, Jansen WJ, Rabinovici GD, et al. Prevalence of Amyloid PET Positivity in Dementia Syndromes: A Meta-analysis HHS Public Access. JAMA 2015;313:1939–49. doi:10.1001/jama.2015.4669

36 Monsell SE, Kukull WA, Roher AE, et al. Characterizing apolipoprotein E ε4 carriers and noncarriers with the clinical diagnosis of mild to moderate Alzheimer dementia and minimal β-amyloid peptide plaques. JAMA Neurol 2015;72:1124–31. doi:10.1001/jamaneurol.2015.1721

37 McKhann GM, Knopman DS, Chertkow H, et al. The diagnosis of dementia due to Alzheimer’s disease: Recommendations from the National Institute on Aging-Alzheimer’s Association workgroups on diagnostic guidelines for Alzheimer’s disease. Alzheimer’s Dement 2011;7:263–9. doi:10.1016/j.jalz.2011.03.005

38 Sabri O, Sabbagh MN, Seibyl J, et al. Florbetaben PET imaging to detect amyloid beta plaques in Alzheimer’s disease: Phase 3 study. Alzheimer’s Dement 2015;11:964–74. doi:10.1016/j.jalz.2015.02.004

39 Rowe CC, Doré V, Jones G, et al. 18F-Florbetaben PET beta-amyloid binding expressed in Centiloids. Eur J Nucl Med Mol Imaging 2017;44:2053–9. doi:10.1007/s00259-017-3749-6

40 Mioshi E, Dawson K, Mitchell J, et al. The Addenbrooke’s Cognitive Examination revised (ACE-R): A brief cognitive test battery for dementia screening. Int J Geriatr Psychiatry 2006;21:1078–85. doi:10.1002/gps.1610

41 Folstein MF, Folstein SE, McHugh PR. ‘Mini-mental state’. A practical method for grading the cognitive state of patients for the clinician. J Psychiatr Res 1975;12:189– 98. doi:10.1016/0022-3956(75)90026-6

42 Hughes CP, Berg L, Danziger WL, et al. A new clinical scale for the staging of dementia. Br J Psychiatry 1982;140:566–72. doi:10.1192/bjp.140.6.566

43 Morris JC. The clinical dementia rating (cdr): Current version and scoring rules. Neurology 1993;43:2412–4. doi:10.1212/wnl.43.11.2412-a

44 Washington University Alzheimer’s Disease Research Center. The CDR® Dementia Staging Instrument. SCALE. 1999;:1. doi:20 May 1999

45 Rosen WG, Terry RD, Fuld PA, et al. Pathological verification of ischemic score in differentiation of dementias. Ann Neurol 1980;7:486–8. doi:10.1002/ana.410070516

46 Ishihara T, Terada S. [Geriatric Depression Scale (GDS)]. Nihon Rinsho 2011;69 Suppl 8:455–8. doi:10.1300/j018v05n01_09

47 Spielberger CD, Gorsuch RL, Lushene RE. Manual for the State-Trait Anxiety Inventory. 1970.

48 Buysse DJ, Reynolds CF, Monk TH, et al. The Pittsburgh sleep quality index: A new instrument for psychiatric practice and research. Psychiatry Res 1989;28:193–213. doi:10.1016/0165-1781(89)90047-4

49 Jutten RJ, Peeters CFW, Leijdesdorff SMJ, et al. Detecting functional decline from normal aging to dementia: Development and validation of a short version of the Amsterdam IADL Questionnaire. Alzheimer’s Dement Diagnosis, Assess Dis Monit 2017;8:26–35. doi:10.1016/j.dadm.2017.03.002

50 Ismail Z, Agüera-Ortiz L, Brodaty H, et al. The Mild Behavioral Impairment Checklist (MBI-C): A Rating Scale for Neuropsychiatric Symptoms in Pre-Dementia Populations. J Alzheimer’s Dis 2017;56:929–38. doi:10.3233/JAD-160979

51 Koychev I, Lawson J, Chessell T, et al. Deep and Frequent Phenotyping study protocol: an observational study in prodromal Alzheimer’s disease. BMJ Open 2019;9:24498. doi:10.1136/bmjopen-2018-024498

52 Solomon A, Kivipelto M, Molinuevo JL, et al. European Prevention of Alzheimer’s Dementia Longitudinal Cohort Study (EPAD LCS): Study protocol. BMJ Open 2018;8. doi:10.1136/bmjopen-2017-021017

53 Ritchie K, Ropacki M, Albala B, et al. Recommended cognitive outcomes in preclinical Alzheimer’s disease: Consensus statement from the European Prevention of Alzheimer’s Dementia project. Alzheimer’s Dement. 2017;13:186–95. doi:10.1016/j.jalz.2016.07.154

54 Donohue MC, Sperling RA, Salmon DP, et al. The preclinical Alzheimer cognitive composite: Measuring amyloid-related decline. JAMA Neurol 2014;71:961–70. doi:10.1001/jamaneurol.2014.803

55 Randolph C, Tierney MC, Mohr E, et al. The Repeatable Battery for the Assessment of Neuropsychological Status (RBANS): Preliminary clinical validity. J Clin Exp Neuropsychol 1998;20:310–9. doi:10.1076/jcen.20.3.310.823

56 Wechsler D. Wechsler Adult Intelligence Scale - Revised. San Antonio, TX: Psychological Corporation 1981.

57 Grober E, Veroff AE, Lipton RB. Temporal unfolding of declining episodic memory on the Free and Cued Selective Reminding Test in the predementia phase of Alzheimer’s disease: Implications for clinical trials. Alzheimer’s Dement Diagnosis, Assess Dis Monit 2018;10:161–71. doi:10.1016/j.dadm.2017.12.004

58 Wechsler D. Wechsler Memory Scale - Third UK Edition. London: Harcourt Assessment 1999.

59 Nelson HE, Willison JR. National Adult Reading Test (NART): Test Manual (2nd edn). 1991.

60 Bowie CR, Harvey PD. Administration and interpretation of the Trail Making Test. Nat Protoc 2006;1:2277–81. doi:10.1038/nprot.2006.390

61 Cambridge Cognition. CANTAB® [Cognitive assessment software]. 2019.

62 Hartley T, Bird CM, Chan D, et al. The hippocampus is required for short-term topographical memory in humans. Hippocampus 2007;17:34–8. doi:10.1002/hipo.20240

63 Kiebel SJ, Garrido MI, Friston KJ. Dynamic causal modelling of evoked responses: The role of intrinsic connections. Neuroimage 2007;36:332–45. doi:10.1016/j.neuroimage.2007.02.046

64 Friston K. A theory of cortical responses. Philos Trans R Soc B Biol Sci 2005;360:815–36. doi:10.1098/rstb.2005.1622

65 De Rover M, Pironti VA, McCabe JA, et al. Hippocampal dysfunction in patients with mild cognitive impairment: A functional neuroimaging study of a visuospatial paired associates learning task. Neuropsychologia 2011;49:2060–70. doi:10.1016/j.neuropsychologia.2011.03.037

66 Blackwell AD, Sahakian BJ, Vesey R, et al. Detecting dementia: Novel neuropsychological markers of preclinical Alzheimer’s disease. Dement Geriatr Cogn Disord 2004;17:42–8. doi:10.1159/000074081

67 Tournier JD, Smith R, Raffelt D, et al. MRtrix3: A fast, flexible and open software framework for medical image processing and visualisation. Neuroimage. 2019;202. doi:10.1016/j.neuroimage.2019.116137

68 American Psychiatric Association. Diagnostic and Statistical Manual of Mental Disorders. Fifth. American Psychiatric Association 2013. doi:10.1176/appi.books.9780890425596

